# BRAZIL IS PROJECTED TO BE THE NEXT GLOBAL COVID-19 PANDEMIC EPICENTER

**DOI:** 10.1101/2020.04.28.20083675

**Authors:** Pedro de Lemos Menezes, David M. Garner, Vitor E. Valenti

## Abstract

Coronavirus disease 2019 (COVID-19) is a disease triggered by SARS-CoV-2 infection, which is related in the most recent pandemic situation, significantly affecting health and economic systems. In this study we assessed the death rate associated to COVID-19 in Brazil and the United States of America (USA) to estimate the probability of Brazil becoming the next pandemic epicenter. We equated data between Brazil and USA obtained through the Worldometer website (www.worldometer.info). Epidemic curves from Brazil and USA were associated and regression analysis was undertaken to predict the Brazilian death rate regarding COVID-19 in June. In view of data from April 9^th^ 2020, death rates in Brazil follow a similar exponential increase to USA (r=0.999; p<0.001), estimating 64,310 deaths by June 9^th^ 2020. In brief, our results demonstrated that Brazil follows an analogous progression of COVID-19 deaths cases when compared to USA, signifying that Brazil could be the next global epicenter of COVID-19. We highlight public strategies to decrease the COVID-19 outbreak.

## INTRODUCTION

The Coronavirus disease 2019 (COVID-19) is a new disease that has developed into a global public health concern as it has rapidly extended worldwide. The transmission of severe acute respiratory syndrome Coronavirus 2 (SARS-CoV-2), initiating COVID-19, has led to this urgent pandemic status (Angelos et al, 2020).

This disease initially appeared in Wuhan, China, which became the first epicenter (Nature, 2020). Next, when the cases were partially controlled in China, Europe became affected and was an added epicenter of COVID-19 (Nature, 2020). At present, the United States of America (USA) is the current pandemic epicenter with in excess of 700,000 COVID-19 cases, consistent with the Worldometer website (www.worldometer.info).

In this scenario, we understand that Brazil has the potential for a considerable increase in deaths related to COVID-19 (www.worldometer.info). We comprehend that Brazil has the likelihood to be the next pandemic epicenter of the disease. So, in this report, we aim to evaluate deaths associated to COVID-19 in Brazil and USA, so as to authenticate any likenesses between the countries.

## METHODS

The mortality data for COVID-19, which occurred in Brazil and the USA, was obtained from the Worldometer website (www.worldometer.info).

Worldometer is managed by an international team of developers, researchers, and volunteers’ with the objective of providing global health statistics available in a challenging and time relevant format, to an audience world-wide. It is published by a small and independent digital media company located in the USA. They have no political, governmental, or corporate affiliations.

Deaths were estimated until the 31^st^ day after the 5^th^ death in both countries.

At first, a link was achieved between COVID-19 mortality in the USA, which is related to the Pandemic development process and is considered as the current pandemic epicenter. Temporal modifications were completed, enabling the comparison of data on the Cartesian x-axis, and the adjustment of the number of deaths for each 10,000,000 inhabitants, which allowed the procurement of data on the Cartesian y-axis.

To relate the mortality results between the two countries, the Bivariate correlation test was enforced and the degree of linear relationship was considered by the Spearman coefficient.

Formerly, there were some regressions to discover the best mathematical model which would allow the representation of the mortality estimation curve for COVID-19 in Brazil and USA. Whilst searching for the mathematical model that best fits the mortality estimation curves, the following models were evaluated: Linear, Logarithmic, Inverse, Quadratic, Cubic, Composite, S, Growth, Exponential and Logistic. For this estimate, a significance of alpha, (α) less than 0.01 (p <0.01, <1%) and R^2^ > 0.95 was adopted. Furthermore, the one-way analysis of variance (ANOVA1) test was applied.

The data were managed via Microsoft Excel version 16.36, for MacOS Catalina version 10.15.4. Statistical assessments were computed by IBM SPSS Statistic Subscription application 1.0.0.1347 64Bits for MacOS. These values were considered statistically significant when (p <0.05, <5%) and the established beta, (β) value was 0.1.

## RESULTS

Until the 31^st^ day after the 5^th^ death, the increase in mortality in Brazil and USA follow a similar pattern. The bivariate correlation test was significant with p <0.001 and degree of linear relationship was R^2^ = 0.99 (Figure 1).

**Figure 1:**
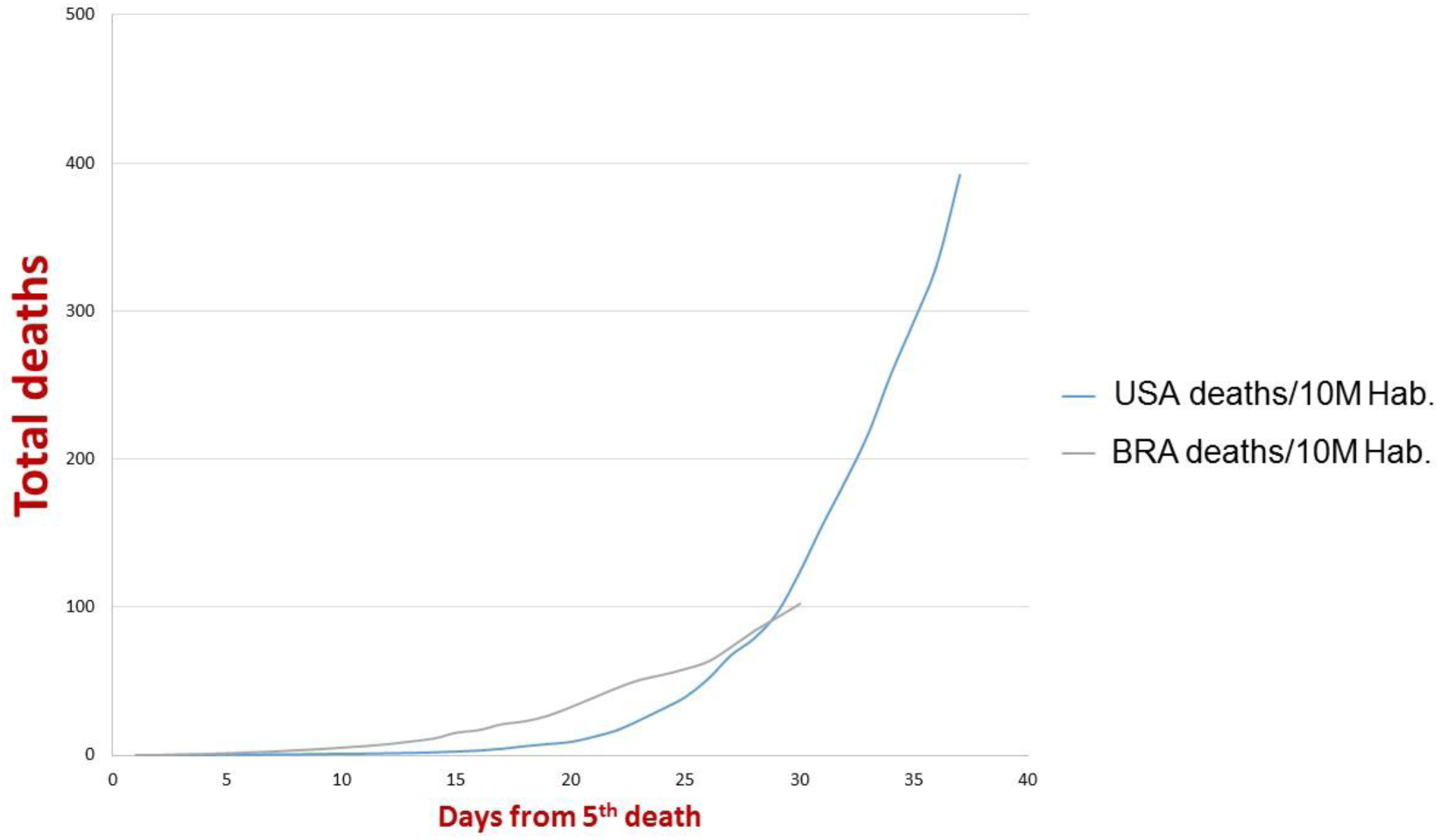
Deaths associated to COVID-19 in Brazil and USA.

In Brazil, the mathematical models attempted to estimate the mortality curve in absolute values, from the 5^th^ to the 30^th^ death (April 17^th^ 2020) and the models that came nearest were the cubic and the quadratic, as illustrated in Figure 2.

**Figure 2:**
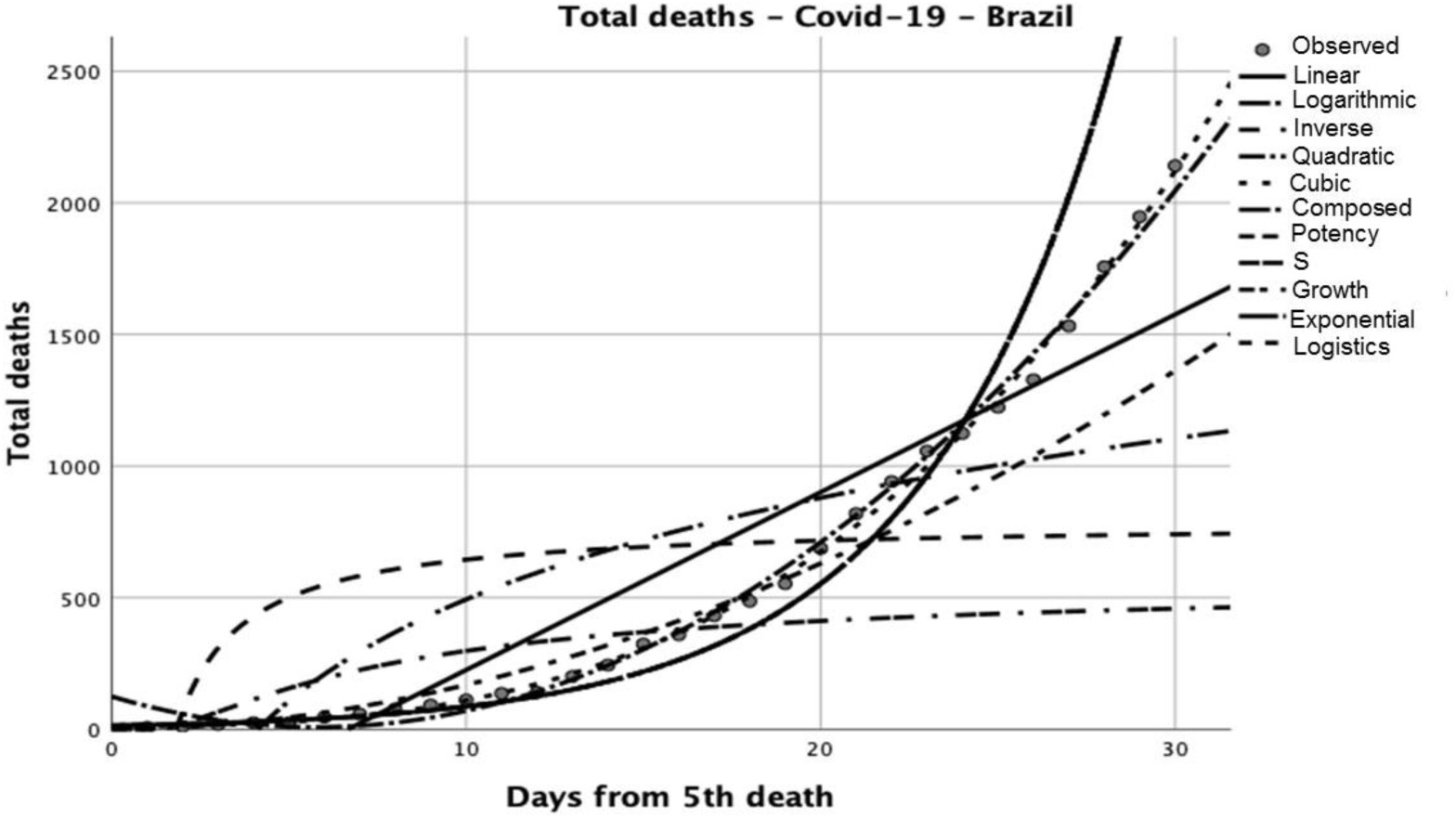
Total deaths related to COVID-19 in Brazil.

Nevertheless, as illustrated in Table 1, the cubic model fits best, R^2^ = 0.998.

Consequently, the cubic model, thus far, best fits an approximation of the growth curve of mortality owing to COVID-19 in Brazil and is in the following manner:

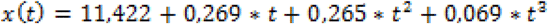

**Table 1:** Model summary and parameter estimates.

| Dependent variable: Deaths |  |  |  |  |  |  |  |  |  |
| --- | --- | --- | --- | --- | --- | --- | --- | --- | --- |
| Equation | R<sup>2</sup> | Model summary |  |  |  | Parameter estimates |  |  |  |
|  |  | Z | df1 | df2 | Sig. | Constant | b1 | b2 | b3 |
| Linear | .859 | 170.682 | 1 | 28 | .000 | -450.108 | 67.532 |  |  |
| Logarithmic | .545 | 33.587 | 1 | 28 | .000 | -789.488 | 556.986 |  |  |
| Invers | .188 | 6.495 | 1 | 28 | .017 | 788.676 | -1442.125 |  |  |
| Quadratic | .995 | 2654.798 | 2 | 27 | .000 | 124.413 | -40.191 | 3.475 |  |
| **Cubic** | **.998** | **4350.630** | **3** | **26** | **.000** | **11.422** | **.269** | **.265** | **.069** |
| Composed | .963 | 732.622 | 1 | 28 | .000 | 13.567 | 1.204 |  |  |
| Potency | .947 | 497.163 | 1 | 28 | .000 | 2.110 | 1.902 |  |  |
| S | .565 | 36.326 | 1 | 28 | .000 | 6.343 | -6.475 |  |  |
| Growth | .963 | 732.622 | 1 | 28 | .000 | 2.608 | .185 |  |  |
| Exponenctial | .963 | 732.622 | 1 | 28 | .000 | 13.567 | .185 |  |  |
| Logistic | .963 | 732.622 | 1 | 28 | .000 | .074 | .831 |  |  |

The independent variable is Days from the 5<sup>th</sup> Death.

Where *t* is the number of days from the 5^th^ death.

With regards this mathematical model, it was demonstrated in Figure 3 that the estimation of total deaths for Brazil, assuming no behavioral changes amongst the population for the 50^th^ day after the 5^th^ death is 9,312 deaths by COVID-19.

**Figure 3:**
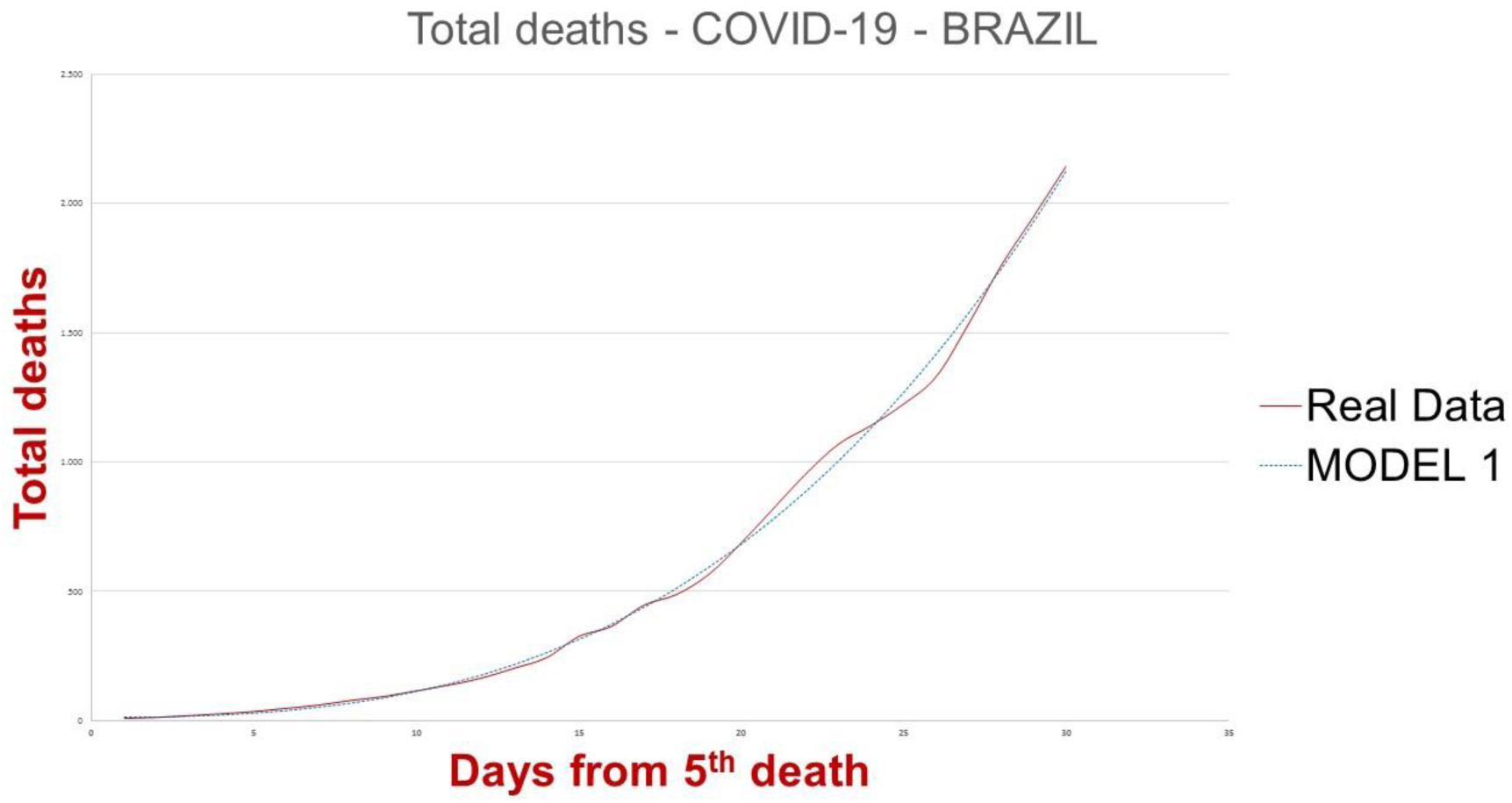
Comparison between total deaths related to COVID-19 in Brazil and projected via mathematical model.

## DISCUSSION

Our brief report was commenced to assess the level of COVID-19 related deaths in Brazil and USA. This intention was explained so as to estimate the possibility of Brazil becoming the next pandemic global epicenter. Our results described significant connections between Brazil and USA about total mortality related to COVID-19 up until the 31^st^ day after the 5^th^ death.

The main concern regarding this mathematical forecast is to enable data to permit the Brazilian government to format policies to protect its own population.

Our study highlights some problems which need addressing. Some Brazilian states, including Sao Paulo, reported only a few COVID-19 cases (Sao Paulo sanctions warnings only in severe cases of COVID-19 and declares that it maintains advice from the Brazilian Ministry of Health), we emphasize the potential of the under reporting of cases.

In summary, the mathematical model enforced in Brazil and USA advocates that Brazil may be the next global pandemic epicenter. We encourage adherence to Brazilian public guidelines to reduce the impact of COVID-19 in Brazil.

## Data Availability

The data are available at www.worldometer.info

## ACKNOWLEDGMENTS

Authors thank the technical staff from Sao Paulo State University, UNESP/Marilia, SP, Brazil and University of Health Sciences of Alagoas, AL, Brazil. Dr. Vitor receives financial support from FAPESP (Process number 2012/01366-6) financial support from the National Council for Scientific and Technological Development, an entity linked to the Ministry of Science, Technology, Innovations and Communications from Brazil (Process number 302197/2018-4).

## AUTHOR CONTRIBUTIONS

Pedro de Lemos Menezes draft the manuscript, wrote introduction and discussion section gave final approval for the version submitted for publication.

David M. Garner draft the manuscript, performed statistical analysis and improved interpretation analysis, reviewed English Grammar and Spelling.

Vitor E. Valenti draft the manuscript, wrote introduction and discussion section and gave final approval for the version submitted for publication.

